# Development, calibration, and external validation of a semi-Markov model for hypertension in older adults with Medicare coverage in the United States

**DOI:** 10.1101/2025.07.02.25330766

**Authors:** Mahip Acharya, Corey J Hayes, Cari A Bogulski, Mir M Ali, Filmon Wolde, Hari Eswaran

## Abstract

**Objective:** To develop a semi-Markov model of hypertension specifically using estimates from primary analysis of Medicare data, and calibrate and validate using external trial and observational data.

**Methods:** A semi-Markov model of hypertension with ten disease states was developed using clinical expertise and literature review: hypertension, myocardial infarction (MI), congestive heart failure (CHF), stroke, transient ischemic attack (TIA), other cardiovascular disease (CVD), chronic CVD, early-stage chronic kidney disease (ES CKD), late-stage CKD (LS CKD), and death. Most transition probabilities and hazard ratios for excess mortality were obtained from analysis of Medicare data (2018–2021). Transition probabilities and hazard ratios were calibrated against findings from the Systolic Hypertension in the Elderly Program (SHEP) trial and post-trial follow-up studies. Iterative grid search using Latin Hypercube Sampling was used for calibration, followed by a recalibration using Nelder-Mead simplex method with common random numbers. External validation was performed using mortality data after MI and stroke.

**Results:** Final monthly transition probabilities from hypertension to other states were: other CVD (0.026), ES CKD (0.010), CHF (0.004), stroke (0.004), TIA (0.002), and MI (0.002). Final hazard ratios were: MI (3.29), CHF (3.99), stroke (3.79), TIA (1.39), other CVD (3.84), chronic CVD (1.39), ES CKD (1.24), and LS CKD (4.16). In the external validation, mortality rates per 100 person-years among stroke survivors were 7.8 (modeled) and 8.2 (observed).

**Conclusion:** Our model was reasonably calibrated except for CHF incidence and stroke-related mortality. External validation showed the model performed well over longer timeframe, but discrepancies were observed for shorter periods.

## Background

Individual- and cohort-level simulation models are used in healthcare decision analysis to understand disease progression and to evaluate cost-effectiveness and budget impacts of preventive and therapeutic interventions.^1–3^ Patients with common or distinct demographic and clinical characteristics are simulated through defined disease states to capture estimated healthcare costs, life-years, quality-adjusted life years, equal value of life years gained, and health years in total.^4–6^ These decision-analytic models are developed using domain knowledge, literature review, and statistical analysis of randomized controlled trials, surveys, insurance claims, or medical records data.^7,8^ Since some input parameters may be unknown or estimated imprecisely given the highly heterogeneous nature of input data, their values are generally assumed and are procedurally adjusted to obtain outputs that match the real-world data (calibration).^9,10^ Different calibration methods exist to calibrate Markov (state-transition cohort) and microsimulation models.^11,12^ The model fit is evaluated using different methods which could be distance-based (sum of squared errors, deviance) or likelihood-based.^13,14^ The model outputs are compared to the real-world estimates—using data different than the ones used for calibration—to evaluate how closely they match (external validation).^15^

Researchers have developed Markov and microsimulation models for hypertension, stroke, and overall cardiovascular disease; ^16–25^ however, these models have several limitations. First, they are constructed, calibrated, and validated in the general population and are not specific to older adults with existing hypertension. ^19,20,23,25^ Second, they use data from years prior to coronavirus disease-19 (COVID-19). ^16–25^ Third, these models are not open source. ^16–25^ Moreover, developing a well-calibrated and validated decision-analytic model for Medicare patients could inform policy-level interventions for the Centers for Medicare & Medicaid Services (CMS). Open source, publicly available, disease simulation models for hypertension could help evaluate public health programs for hypertension control, assess patient-level interventions for management and treatment of hypertension and comorbid cardiovascular diseases, and work as a foundation for the development of more nuanced and bespoke hypertension and cardiovascular disease models.

The objectives of this study were to develop, calibrate, and validate a microsimulation model with semi-Markov properties for hypertensive patients using an open source environment (R/RStudio). This model was developed to assess the cost-effectiveness and budget impact of remote patient monitoring for hypertension management in patients aged 65 or older with traditional Medicare coverage.

## Methods

### Model development

We developed a semi-Markov model for individual-level (microsimulation) and cohort-level simulation of older adults with hypertension. We developed the model using our clinical expertise, existing clinical literature, and published decision analytic models.^16,20–22,26,27^ The model included ten disease states: hypertension, myocardial infarction (MI), congestive heart failure (CHF), stroke, transient ischemic attack (TIA), other cardiovascular disease (CVD), chronic CVD, early-stage chronic kidney disease (ES CKD), late-stage chronic kidney disease (LS CKD), and death (Figure 1). MI, CHF, stroke, TIA, and other CVD states were considered acute CVD states. All patients started in the hypertension state and could transition to one of the acute CVD states, the ES CKD state, or death (from non-cardiovascular causes). Twelve tunnel states were used for each of the five acute CVD states to model higher risk of death (cardiovascular-related). After transitioning to any of the acute CVD states and subsequently through the tunnel states, patients would enter the chronic CVD state. Patients could also transition from hypertension to ES CKD, from which they could transition to the LS CKD state or death. Patients could not transition back to the hypertension state from any other state. To construct a relatively simpler model, no transitions were allowed between cardiovascular disease states and the CKD states.

**Figure 1:**
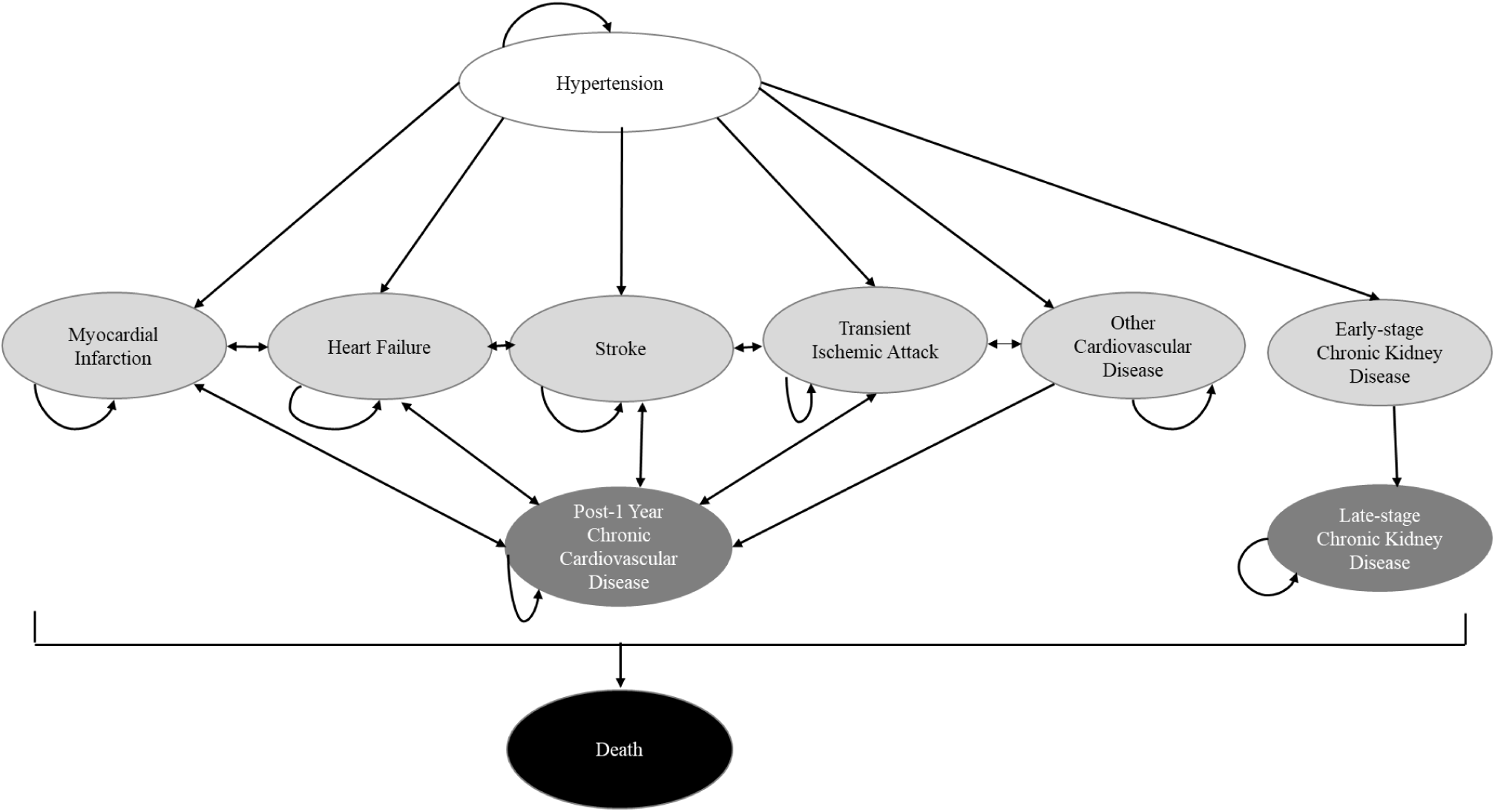
Semi-Markov Model for Hypertension.

The transition probabilities from hypertension to acute CVD states and the ES CKD state were derived from the analysis of a 20% random sample of Medicare Fee-For-Service database (2018–2021). First, a cohort of individuals diagnosed with hypertension in an outpatient setting were identified using International Classification of Diseases-10-Clinical Modification (ICD-10-CM) diagnosis codes. The first hypertension diagnosis between July 1, 2018 and June 30, 2020 was determined and considered the index date. Individuals were required to have continuous coverage for Medicare parts A and B in the six months prior to the index date (baseline period). To consider only incident hypertensive patients in the analysis, patients were required to have no outpatient hypertension diagnosis in the baseline period. The sample was further restricted to patients aged 65–100 years on index date. The patients in the final analytical sample were then followed for a maximum of one year to determine the incident diagnosis of MI, CHF, stroke, TIA, other CVD, and ES CKD. These conditions were identified using ICD-10-CM diagnosis codes. Other CVD included arrhythmias, valvular heart disease, and peripheral artery disease. Kaplan-Meier survival rates were estimated for the six conditions (reflecting the six disease states in the microsimulation model), with death prior to the event of interest as a competing event. These analyses gave us annual transition rates from hypertension to the six disease states.

To estimate the transition rates from MI, CHF, stroke, TIA, and other CVD to each other (after passing through the tunnel states), patients with each of the five conditions in the one-year follow-up were identified and included. For example, patients who had an incident MI diagnosis in the follow-up were identified (MI cohort); the first MI diagnosis date was considered as the new index date; and the patients were followed from this new index date for a maximum of one year or until an incident diagnosis for CHF, stroke, TIA, or other CVD. Kaplan-Meier survival rates were calculated using death as a competing event. These analyses resulted in annual transition rates from the five acute CVD states to each acute CVD state except for itself. We used a slightly different approach to estimate transition rates for the recurrence of acute CVD events (for example, from MI to subsequent MI), as a subsequent diagnosis code for the same condition may not necessarily be a new acute event and could be a documentation of the patient’s event history. To identify the subsequent event, we only considered the primary diagnosis codes and only those documented in an emergency department (ED) visit or a hospitalization. Kaplan-Meier estimates with death as a competing event were calculated the same way as described above. These analyses yielded annual transition rates from MI to MI, CHF to CHF, stroke to stroke, TIA to TIA, and other CVD to other CVD. Although the recurrences of CHF or other CVD may not reflect the occurrence of new CHF event (as they could be considered chronic conditions), we still modeled them as they capture a hospitalization or ED visit associated with that event and thereby have significance for healthcare costs and mortality. The annual transition rates from chronic CVD to the acute CVD events were also modeled, as patients could have future events, and the rates were assumed to be the same as that from the other CVD state to remaining acute CVD states.

We also calculated excess mortality in one year following each of the acute CVD events. For example, for the MI cohort, we gathered clinical and demographic characteristics from the six-month baseline period: age on index date, sex, race/ethnicity, index year, alcohol use disorder, substance use disorder, depressive disorders, psychosis, neurological disorders, anemia, blood loss disorders, weight loss, diabetes mellitus (uncomplicated), diabetes mellitus (complicated), complications of hypertension, HIV/AIDS, hypothyroidism, liver disease, pneumonia, septicemia, obesity, paraplegic conditions, solid tumors, lymphoma, metastasis, and cardiovascular diseases. These characteristics were selected based on a previous study we conducted using Medicare data to assess mortality .^28^ A non-MI cohort was also identified from the initial hypertensive cohort and the same baseline characteristics as described above were assessed. The MI and non-MI cohorts were then matched on propensity scores that were estimated using the baseline covariates. The matched groups were followed for a maximum of one year or until the patients died. Cox proportional hazard regressions were used to estimate the hazard ratio of MI diagnosis on mortality. The same approach was used to determine hazard ratios for mortality from CHF, stroke, TIA, and other CVD diagnosis. The hazard ratio for mortality following ES CKD was obtained from a study by Fox et al.^29^ LS CKD-associated hazard ratio for mortality was also estimated from the Medicare data, calculated similarly as that for acute CVD events. Background mortality was used from the 2021 life tables published by the Centers for Disease Control and Prevention (CDC).^30^

### Model calibration

#### Calibration parameters

Select transition probabilities, hazard ratios, non-cardiovascular deaths, and cardiovascular deaths were used as calibration parameters. The following transition probabilities were adjusted in the calibration: hypertension to MI, CHF, stroke, TIA, and other CVD; MI to subsequent MI, CHF, stroke, TIA, and other CVD; CHF to MI, subsequent CHF, stroke, TIA and, other CVD; stroke to MI, CHF, subsequent stroke, TIA, and other CVD; TIA to MI, CHF, stroke, subsequent TIA, and other CVD; and chronic CVD to MI, CHF, stroke, and TIA. The transition probabilities were adjusted by ±20% of their initial values in the calibration process. The following hazard ratios for mortality were used as calibration parameters: MI, CHF, stroke, TIA, and other CVD. A common multiplier was used for the five hazard ratios (hazard ratio multiplier). The multiplier ranged from 1.0 to 1.4 in the calibration process. We used the lower bound of 1.0 for the multiplier, since a trial run of the model showed an underestimation of stroke-related and cardiovascular mortality. Another multiplier was used for the share of non-cardiovascular deaths of total deaths, with the CDC life table mortality used as the initial values (mortality multiplier). The values for the mortality multiplier ranged from 0.33 (one-third) to 0.75 (three-fourth) in the calibration process.

#### Calibration targets

The following parameters were used as calibration targets: all-cause mortality, cardiovascular mortality, non-cardiovascular mortality, stroke-related mortality, stroke incidence, TIA incidence, MI incidence, and CHF incidence. Published estimates from the Systolic Hypertension in the Elderly Program (SHEP) trial and the observational follow-up of the trial participants were used.^31–34^ The SHEP trial was a randomized, placebo-controlled, double-blind trial conducted in adults aged 60 or above with isolated systolic hypertension.^35^ The trial had an average follow-up of 4.5 years and its objective was to determine the efficacy of antihypertensive medications in reducing blood pressure and incidence of stroke.^31^ The trial reported stroke incidence and absolute number of cases of MI, TIA, and CHF for both placebo and treatment groups.^31,32^ We used estimates from the placebo group as our calibration targets. We derived estimates for MI, TIA, and CHF incidences using the number of cases for the respective event, number of stroke events, and the stroke incidence in the placebo group. The three mortality estimates were obtained from the longer follow-up study of SHEP trial participants.^33,34^ The Kaplan-Meier curves for all-cause mortality and cardiovascular mortality were digitized to obtain the mortality incidences. We ran a microsimulation model using the age distribution from the SHEP trial to better align the modeling strategy with the trial and the post-trial follow-up results.

#### Calibration method

We used an empirical calibration approach to determine the input parameter values. An empirical calibration approach entails a data-driven process wherein the models are supplied with varying input values, and the generated outputs are compared to observed data.^12^ Latin Hypercube sampling (LHS)—a grid-based sampling strategy—was used to select the values from the multi-dimensional calibration parameters space.^9^ A total of 1,000 samples of 100,000 individuals were used to ensure that multiple combinations of input parameters were selected and tested. Poisson deviance was computed to determine the goodness of fit, as deviance-based goodness of fit measures are shown to lead to more accurate estimation of the parameters.^14^ As we had eight calibration targets and eight resulting deviance measures, we combined them to one score using equal weighting. We selected the sample and subsequently the model inputs that yielded the lowest deviance value.

After the LHS-based calibration, we reviewed the simulated values for calibration targets and noticed that the model under-estimated stroke-related mortality and over-estimated the CHF incidence. So, we performed a second calibration (recalibration) using the best-fitting inputs from the first calibration and varying the following parameters: hazard ratio of excess mortality from stroke, and transition probabilities to CHF from hypertension, MI, CHF, stroke, TIA, other CVD, and chronic CVD. Nelder-Mead simplex algorithm, an iterative search strategy, was used for the recalibration.^14,36^ We utilized the common random number technique to reduce noise resulting from the stochastic nature of the microsimulation model.^37,38^ Poisson deviance was used as the goodness of fit measure, similar to the previous step, and the same calibration targets were used. The model inputs corresponding to the best-fitting model were selected as the final values.

### External validation

We performed an external validation of the model by using data on mortality rates following MI and stroke events. We obtained the validation targets from two published studies: i) an observational study conducted using an MI registry linked to Medicare data,^39^ and ii) a prospective cohort study conducted among patients who suffered a stroke event and survived at least 30 days.^40^ The following five parameters were extracted: mortality rates per 100, 500, and 800 person-years after an MI event; mortality rates per 800 person-years after one year post-MI event; and the mortality rate per 100 person-years after one month post-stroke event over 12 years of follow-up. The rates from our recalibrated semi-Markov model were compared qualitatively to the reported estimates from the two studies. All relevant codes and data used in the development, calibration, and validation are shared in the GitHub repository.^41^ The study was deemed Not Human Subject Research by University of Arkansas for Medical Sciences Institutional Review Board (#276216).

## Results

All monthly transition probabilities and hazard ratios are listed in Table 1. The probability of transition from the hypertension state was highest to the other CVD state, followed by that to ES CKD, CHF, stroke, TIA, and MI. Based on our analysis of Medicare data, the hazard ratios of MI, CHF, and stroke for all-cause mortality were 3.19, 3.87, and 2.30, respectively (Table 1). The hazard of mortality was the highest after LS CKD (4.03), while it was the lowest following a TIA event (1.35). The monthly probabilities of recurrence of MI, CHF, stroke, and TIA were, respectively, 0.008, 0.010, 0.006, and 0.003.

**Table 1:** Input parameters used in the calibration process.

| **Model Parameters** | **Estimates** | **Source** | **Range used in 1<sup>st</sup> calibration** |
| --- | --- | --- | --- |
| ***Transition Probabilities*** | ***(Monthly)*** |  |  |
| Hypertension to MI | 0.002164 | Primary analysis of Medicare Data (Medicare) | [0.001731, 0.002597] |
| Hypertension to CHF | 0.007422 | Medicare | [0.005938, 0.008907] |
| Hypertension to Stroke | 0.003270 | Medicare | [0.002616, 0.003924] |
| Hypertension to TIA | 0.002231 | Medicare | [0.001785, 0.002677] |
| Hypertension to Other CVD | 0.02574 | Medicare | [0.02059, 0.03089] |
| Hypertension to ES CKD | 0.01039 | Medicare | Fixed |
| MI to MI | 0.008323 | Medicare | [0.006659, 0.009988] |
| MI to CHF | 0.005883 | Medicare | [0.004706, 0.007059] |
| MI to Stroke | 0.002322 | Medicare | [0.001858, 0.002787] |
| MI to TIA | 0.001349 | Medicare | [0.001079, 0.001619] |
| MI to Other CVD | 0.01655 | Medicare | [0.01324, 0.01986] |
| CHF to CHF | 0.01034 | Medicare | [0.008270, 0.01241] |
| CHF to MI | 0.001416 | Medicare | [0.001132, 0.001699] |
| CHF to Stroke | 0.001482 | Medicare | [0.001186, 0.001779] |
| CHF to TIA | 0.0008496 | Medicare | [0.0006797, 0.001020] |
| CHF to Other CVD | 0.01451 | Medicare | [0.01161, 0.01741] |
| Stroke to Stroke | 0.006421 | Medicare | [0.005137, 0.007705] |
| Stroke to MI | 0.001382 | Medicare | [0.001106, 0.001659] |
| Stroke to CHF | 0.003843 | Medicare | [0.003074, 0.004611] |
| Stroke to TIA | 0.002164 | Medicare | [0.001731, 0.002597] |
| Stroke to Other CVD | 0.01073 | Medicare | [0.008587, 0.01288] |
| TIA to TIA | 0.003344 | Medicare | [0.002676, 0.004013] |
| TIA to MI | 0.001258 | Medicare | [0.001006, 0.001509] |
| TIA to CHF | 0.003502 | Medicare | [0.002802, 0.004203] |
| TIA to Stroke | 0.003037 | Medicare | [0.002430, 0.003644] |
| TIA to Other CVD | 0.01105 | Medicare | [0.008837, 0.01326] |
| Other CVD to Other CVD | 0.01920 | Medicare | [0.01536, 0.02305] |
| Other CVD to MI | 0.0005757 | Medicare | [0.0004605, 0.0006908] |
| Other CVD to CHF | 0.002023 | Medicare | [0.001618, 0.002428] |
| Other CVD to Stroke | 0.0007780 | Medicare | [0.0006224, 0.0009336] |
| Other CVD to TIA | 0.0005224 | Medicare | [0.0004179, 0.0006268] |
| Chronic CVD to MI | 0.0005757 | Medicare | [0.0004605, 0.0006908] |
| Chronic CVD to CHF | 0.002023 | Medicare | [0.001618, 0.002428] |
| Chronic CVD to Stroke | 0.0007780 | Medicare | [0.0006224, 0.0009336] |
| Chronic CVD to TIA | 0.0005224 | Medicare | [0.0004179, 0.0006268] |
| ES CKD to LS CKD | 0.002148 | Vupputuri et. al<sup>52</sup> | Fixed |
| **Hazard Ratio Multiplier** |  | Range assumed | [1.00, 1.40] |
| **Hazard Ratios for excess mortality** |  |  |  |
| MI | 3.19 | Medicare | [3.19, 4.47] |
| CHF | 3.87 | Medicare | [3.87, 5.42] |
| Stroke | 2.30 | Medicare | [2.30, 3.22] |
| TIA | 1.35 | Medicare | [1.35, 1.82] |
| Other CVD | 3.72 | Medicare | [3.72, 5.21] |
| Chronic CVD | 1.35 | Assumed similar to that for TIA | [1.35, 1.82] |
| ES CKD | 1.20 | Fox et al.<sup>29</sup> | Fixed |
| ES CKD | 4.03 | Medicare | Fixed |
| **Multiplier for non-cardiovascular background mortality** |  | Range assumed | [0.33, 0.75] |

MI: Myocardial Infarction; CHF: Congestive Heart Failure; TIA: Transient Ischemic Attack; CVD: Cardiovascular Disease; ES CKD: Early-stage Chronic Kidney Disease; LS CKD: Late-stage Chronic Kidney Disease
No transition was allowed from Chronic CVD to the Other CVD state

Calibration results from the best-fitting model after the first round of calibration are shown in Table 2. The modeled (observed) incidences of MI, stroke, and TIA per 100,000 after the first calibration were, respectively: 5,682 (5,054), 8,487 (8,200), and 4,449 (4,229). The model over-estimated CHF incidence and underestimated stroke-related mortality: the modeled CHF incidence and stroke-related mortality, respectively, were14,718 and 1,487, while the corresponding observed values were 5,483 and 5,000.

**Table 2:** Calibration targets and the simulated values in first and second rounds of calibration.

| Calibration targets | Target values (per 100,000 individuals) | Simulated values after 1<sup>st</sup> calibration | Simulated values after 2<sup>nd</sup> calibration (recalibration) |
| --- | --- | --- | --- |
| MI incidence in 5 years | 5,054 | 5,682 | 5,711 |
| CHF incidence in 5 years | 5,483 | 14,718 | 10,766 |
| Stroke incidence in 5 years | 8,200 | 8,487 | 8,518 |
| TIA incidence in 5 years | 4,229 | 4,449 | 4,592 |
| Stroke-related mortality in 22 years | 5,000 | 1,487 | 2,544 |
| Non-cardiovascular mortality in 14 years | 20,700 | 22,590 | 22,846 |
| Cardiovascular mortality in 14 years | 24,300 | 24,058 | 23,869 |
| All-cause mortality in 14 years | 45,000 | 46,648 | 46,715 |

MI: Myocardial Infarction; CHF: Congestive Heart Failure; TIA: Transient Ischemic Attack

Table 3 lists the parameters used in the recalibration and the final parameter values. The final hazard ratio of mortality after a stroke event was 3.79, which was substantially higher than the initial (2.30) and the best-fitting value from the first calibration (2.37). In contrast, the monthly transition probability of hypertension to CHF in the final model was 0.004, which was lower than the initial (0.007) and the value resulting from the first calibration (0.006). The modeled (observed) all-cause, cardiovascular, and non-cardiovascular deaths after the recalibration, respectively, were: 46,715 (45,000), 23,869 (24,300), and 22,590 (20,700) (Table 2). The modeled cumulative incidences (per 100,000) of MI, CHF, stroke, and TIA over 10 years were 7,880, 14,782, 11,261, and 6,460, respectively (Figure 2).

**Figure 2:**
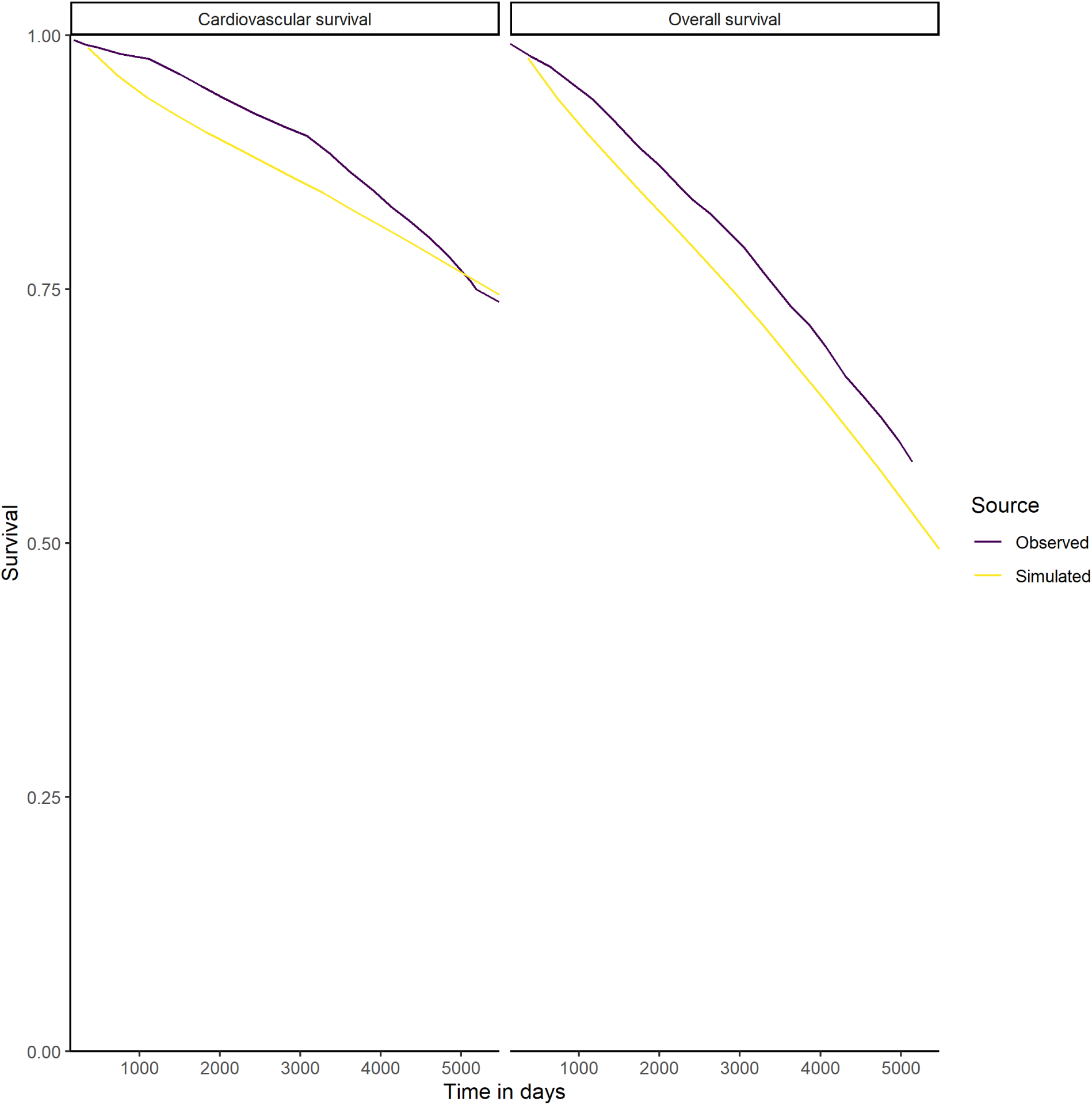
Observed and simulated cardiovascular and overall survival rates after the second round of calibration (recalibration)

**Table 3:** Calibration parameters resulting from best-fitting model in the first round of calibration and used for the second round of calibration (recalibration)

| Parameters | Values | Range in recalibration | Final values |
| --- | --- | --- | --- |
| *Transition Probabilities* |  |  |  |
| Hypertension to MI | 0.002320 | Fixed | 0.002320 |
| Hypertension to CHF | 0.006314 | [0.004420, 0.005683] | 0.004420 |
| Hypertension to Stroke | 0.003726 | Fixed | 0.003726 |
| Hypertension to TIA | 0.001850 | Fixed | 0.001850 |
| Hypertension to Other CVD | 0.02641 | Fixed | 0.02641 |
| Hypertension to ES CKD | 0.01039 | Fixed | 0.01039 |
| MI to MI | 0.008037 | Fixed | 0.008037 |
| MI to CHF | 0.006630 | [0.004641, 0.005967] | 0.005143 |
| MI to Stroke | 0.002442 | Fixed | 0.002442 |
| MI to TIA | 0.001108 | Fixed | 0.001108 |
| MI to Other CVD | 0.01756 | Fixed | 0.01756 |
| CHF to CHF | 0.01120 | [0.007840, 0.01008] | 0.01008 |
| CHF to MI | 0.001233 | Fixed | 0.001233 |
| CHF to Stroke | 0.001385 | Fixed | 0.001385 |
| CHF to TIA | 0.0008625 | Fixed | 0.0008625 |
| CHF to Other CVD | 0.01666 | Fixed | 0.01666 |
| Stroke to Stroke | 0.006269 | Fixed | 0.006269 |
| Stroke to MI | 0.001528 | Fixed | 0.001528 |
| Stroke to CHF | 0.003490 | [0.002443, 0.003141] | 0.002443 |
| Stroke to TIA | 0.001846 | Fixed | 0.001846 |
| Stroke to Other CVD | 0.01122 | Fixed | 0.01122 |
| TIA to TIA | 0.003241 | Fixed | 0.003241 |
| TIA to MI | 0.001440 | Fixed | 0.001440 |
| TIA to CHF | 0.003757 | [0.002630, 0.003881] | 0.003381 |
| TIA to Stroke | 0.002513 | Fixed | 0.002513 |
| TIA to Other CVD | 0.01014 | Fixed | 0.01014 |
| Other CVD to Other CVD | 0.02260 | Fixed | 0.02260 |
| Other CVD to MI | 0.0006741 | Fixed | 0.0006741 |
| Other CVD to CHF | 0.001958 | [0.001371, 0.001762] | 0.001371 |
| Other CVD to Stroke | 0.0008530 | Fixed | 0.0008530 |
| Other CVD to TIA | 0.0005698 | Fixed | 0.0005698 |
| Chronic CVD to MI | 0.0006780 | Fixed | 0.0006780 |
| Chronic CVD to CHF | 0.001876 | [0.001313, 0.001688] | 0.001313 |
| Chronic CVD to Stroke | 0.0008825 | Fixed | 0.0008825 |
| Chronic CVD to TIA | 0.0005722 | Fixed | 0.0005722 |
| ES CKD to LS CKD | 0.002148 | Fixed | 0.002148 |
| ***Hazard Ratios for excess mortality*** |  |  |  |
| MI | 3.29 | Fixed | 3.29 |
| CHF | 3.99 | Fixed | 3.99 |
| Stroke | 2.37 | [2.84, 3.79] | 3.79 |
| TIA | 1.39 | Fixed | 1.39 |
| Other CVD | 3.84 | Fixed | 3.84 |
| Chronic CVD | 1.39 | Fixed | 1.39 |
| ES CKD | 1.24 | Fixed | 1.24 |
| LS CKD | 4.16 | Fixed | 4.16 |
| ***Multiplier for non-cardiovascular background mortality*** | 0.3465 | Fixed | 0.3465 |

MI: Myocardial Infarction; CHF: Congestive Heart Failure; TIA: Transient Ischemic Attack; CVD: Cardiovascular Disease; ES CKD: Early-stage Chronic Kidney Disease

In the external validation of the recalibrated model, the modeled (observed) mortality rates per 100, 500, 800 person-years following an MI event were 15.5 (24.0), 41.0 (51.0), and 61.7 (65.0) (Figure 3). Among the individuals who survived one year after an MI event, the modeled mortality rate per 800 person-years was 50.1 compared to the observed rate of 59.1. The modeled and observed mortality rates among individuals with stroke who survived for at least 30 days were similar (7.8 versus 8.2, respectively).

**Figure 3:**
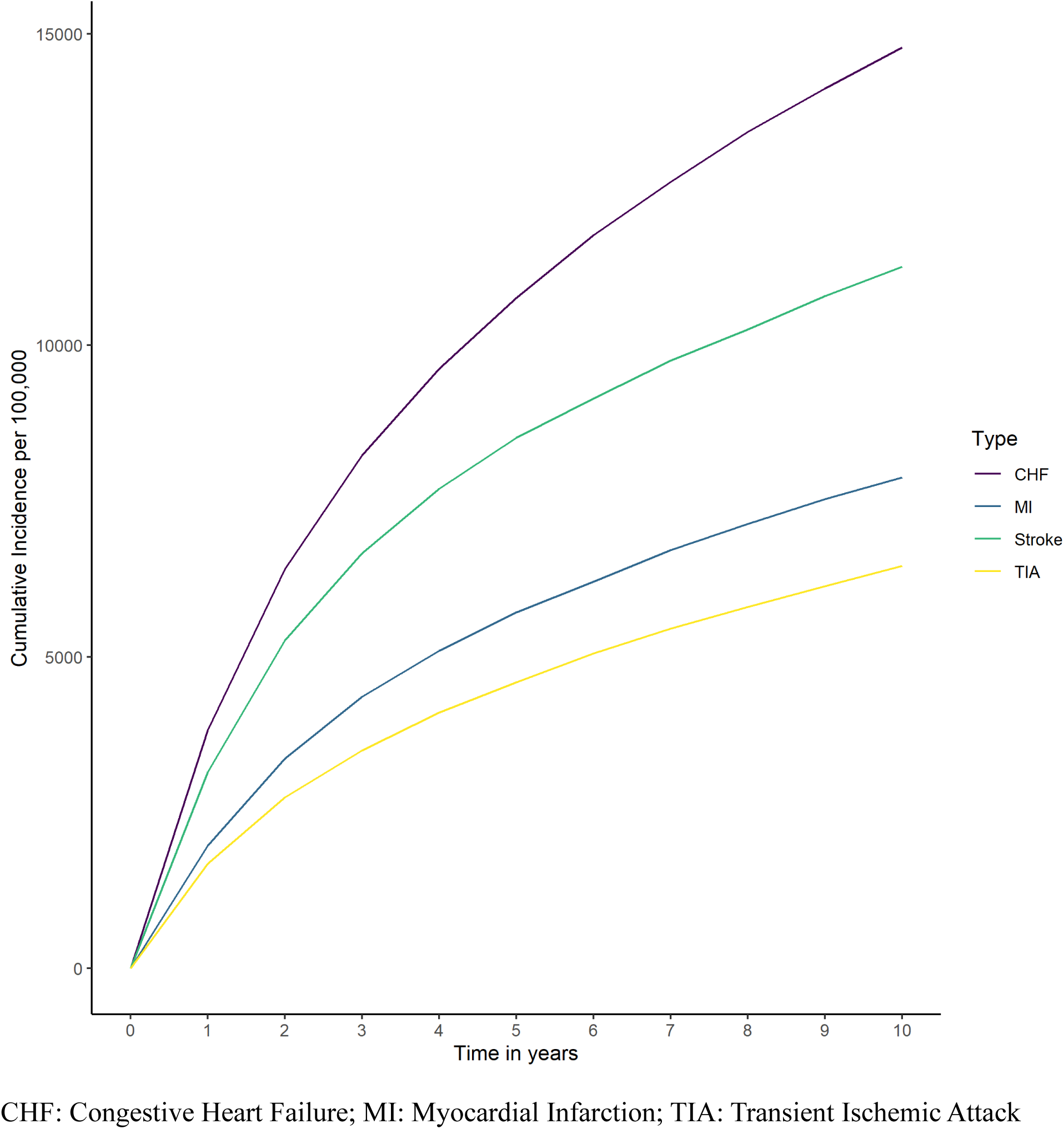
Simulated cumulative incidences of MI, CHF, Stroke and TIA per 100,000 after the second round of calibration (recalibration)

**Figure 4:**
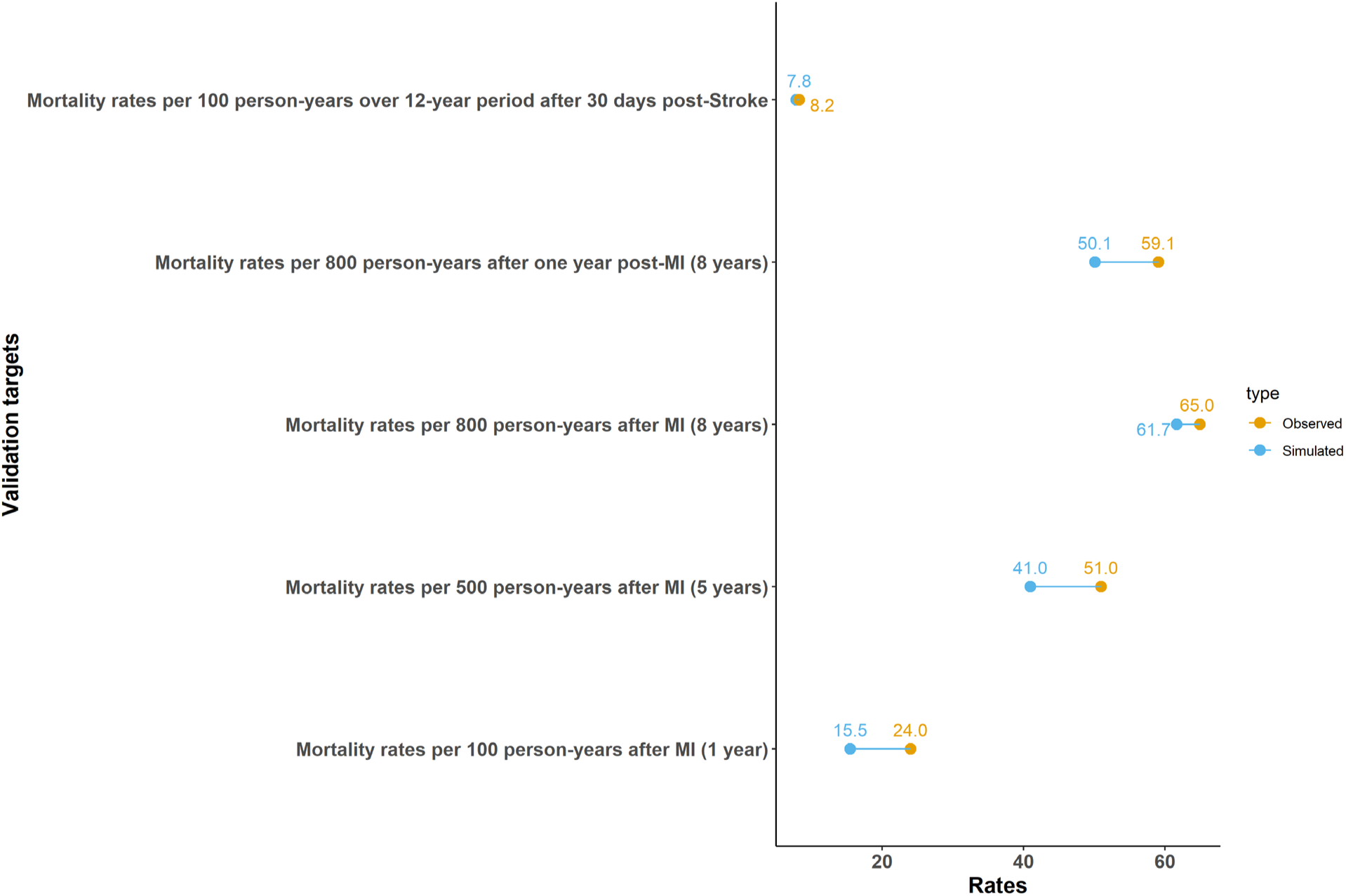
External validation results of the microsimulation model.

## Discussion

In this study, we developed, calibrated, and validated a microsimulation model with semi-Markov property for hypertension among older adults with Medicare. Based on the qualitative comparison, the calibrated model displayed a good fit to the data from the SHEP trial for mortality measures and incidences of MI, stroke, and TIA.^31–34^ However, the model under-estimated stroke-related mortality, and over-estimated the CHF incidence. The SHEP trial started their enrollment in 1985,^35^ while the input parameters for this study were derived from Medicare 2018–2021, so some of the discrepancies between our modeled and observed rates could be attributed to the difference in the timeframe reflecting the changes in hypertension management and treatment practices.

The onset of the COVID-19 public health emergency (PHE) led to a reduction in dispensing of antihypertensive medications, which has been predicted to increase acute CVD events such as MI and stroke.^42^ On the other hand, stroke-related deaths have been steadily declining owing to the advances in early identification and management.^43^ Our modeled estimates appear to align with these findings, particularly with respect to a higher incidence of acute CVD events and lower stroke-related mortality. Additionally, the CDC mortality tables reveal an increase in age-adjusted mortality from 2019 through 2021, ^30,44,45^ which could have led to the slight over-estimation of mortality in our simulation compared to the trial data. As a result, we also limited the range of input parameters used in calibration to avoid overfitting the model to the older calibration data. Going forward, observational and/or trial data are needed for the rates of CVD and CKD estimated during the COVID-19 PHE. In the external validation, the modeled estimates were closer to the observed values when the follow-up time was longer, indicating that the model may be mis-calibrated over a shorter timeframe and time-varying probabilities and hazard ratios, at a more granular level, may be required. Despite the discrepancy between the observed and modeled estimates, the open-source codes of our model would make it easier to re-calibrate against a different set of calibration targets.

Following the onset of the COVID-19 PHE, the use of digital health technologies and virtual visits skyrocketed in 2020, and although utilization rates have declined, they remain higher than rates observed prior to the PHE.^46,47^ Chronic disease management also had a seismic shift, with increased adoption of remote patient and therapeutic monitoring,^48,49^ which involve collection of patient data without direct interaction with a clinician and the subsequent storage and transfer of these data via virtual connections such as Bluetooth and Wi-Fi.^50^ Given the increased use of these technologies, and more so in certain demographics (older adults), models and the model input parameters used for evaluating their effectiveness and cost-effectiveness need to reflect the current disease progression rates and the healthcare utilization paradigm. The development, calibration and external validation of this hypertension disease model was motivated by our ongoing evaluation of effectiveness, cost-effectiveness, and budget impact of remote patient monitoring for hypertension management in Medicare enrollees aged 65 years and older.

Our study has several limitations. First, we estimated transition probabilities using ICD-10-CM diagnosis codes in the Medicare claims database, which are used for billing purposes and may not accurately capture incident events. Although we used a six-month washout period to establish incidence, it may not be adequate, and the diagnosis codes may reflect history of the event. Second, we used empirical calibration for the first calibration, which does not have a closed-form solution and may not yield the best fit value for each calibrated parameter. As a result, the transition probabilities in our study should not be used in isolation. Additionally, going forward, if there are changes in the hypertension management paradigm, then the model may need to be recalibrated. Third, the calibration and validation data themselves could suffer from issues such as selection bias and measurement error and may deviate from the true cardiovascular event rates. For example, the proportions of cardiovascular and non-cardiovascular mortality in the SHEP trial appear to be substantially different from those observed in the Systolic Blood Pressure Intervention Trial (SPRINT) study.^51^ These discrepancies could likely be attributed to differences in care practice in the enrollment sites, patient population (different comorbidity profile, healthcare utilization patterns), and/or random variation.

In conclusion, we developed an open-source semi-Markov model for hypertension in older adults, calibrated it using individual-level simulation against a published trial and post-trial follow-up data, and externally validated the results against outputs from two different data sources. The model was reasonably calibrated for incidences of acute CVD events except for CHF and for mortality measures except for stroke-related mortality. This developed model can be used for evaluation of hypertension-related interventions in the aftermath of the COVID-19 pandemic, although recalibrating it when more recent and better-quality data become available would improve it.

## Data Availability

The aggregate data used in the study are posted on the GitHub repository at https://github.com/AacharyaM/Hypertension-modeling

---

**Supplement Table 1:** Identification of clinical characteristics using diagnoses.

| **Characteristics** | **Claims-based identification** |
| --- | --- |
| Hypertension | ICD-10-CM: 'I10' |
| Hypertension, complicated | ICD-10-CM: 'I11', 'I12', 'I13', 'I15' |
| Any ED visit | CPT: '99281', '99282', '99283', '99284', '99285', '99291', '99292', '99234', '99235', '99236', '99217', '99218', '99219', '99220', '99224', '99225', '99226' AND<br>Place of service: '23' |
| Myocardial infarction | ICD-10-CM: 'I21', 'I22' |
| Congestive heart failure | ICD-10-CM: 'I099', 'I110', 'I130', 'I132', 'I255', 'I420', 'I425', 'I426', 'I427', 'I428', 'I429', 'I43', 'I50', 'P290' |
| Cardiac arrhythmia | ICD-10-CM: 'I47', 'I48', 'I49', 'I441', 'I442', 'I443', 'I456', 'I459', 'R000', 'R001', 'R008', 'T821', 'Z450', 'Z950' |
| Valvular disease | ICD-10-CM: 'I70', 'I71', 'I731', 'I738', 'I739', 'I771', 'I790', 'I792', 'K551', 'K558', 'K559', 'Z958', 'Z959' |
| Circulation disorders | ICD-10-CM: 'I26', 'I27', 'I280', 'I288', 'I289' |
| Peripheral vascular disorders | ICD-10-CM: 'I70', 'I71', 'I731', 'I738', 'I739', 'I771', 'I790', 'I792', 'K551', 'K558', 'K559', 'Z958', 'Z959' |
| Paralysis | ICD-10-CM: 'G81', 'G82', 'G041', 'G114', 'G801', 'G802', 'G830', 'G831', 'G832', 'G833', 'G834', 'G839' |
| Other Neurological disorders | ICD-10-CM: 'G10', 'G11', 'G12', 'G13', 'G20', 'G21', 'G22', 'G32', 'G35', 'G36', 'G37', 'G40', 'G41', 'R56', 'G254', 'G255', 'G312', 'G318', 'G319', 'G931', 'G934', 'R470' |
| Chronic pulmonary disease | ICD-10-CM: 'I278', 'I279', 'J684', 'J701', 'J703', 'J40', 'J41', 'J42', 'J43', 'J44', 'J45', 'J46', 'J47', 'J60', 'J61', 'J62', 'J63', 'J64', 'J65', 'J66', 'J67' |
| Diabetes, uncomplicated | ICD-10-CM: 'E100', 'E101', 'E109', 'E110', 'E111', 'E119', 'E120', 'E121', 'E129', 'E130', 'E131', 'E139', 'E140', 'E141', 'E149' |
| Diabetes, complicated | ICD-10-CM: 'E102', 'E103', 'E104', 'E105', 'E106', 'E107', 'E108', 'E112', 'E113', 'E114', 'E115', 'E116', 'E117', 'E118', 'E122', 'E123', 'E124', 'E125', 'E126', 'E127', 'E128', 'E132', 'E133', 'E134', 'E135', 'E136', 'E137', 'E138', 'E142', 'E143', 'E144', 'E145', 'E146', 'E147', 'E148' |
| Hypothyroidism | ICD-10-CM: 'E00', 'E01', 'E02', 'E03', 'E890' |
| Renal failure | ICD-10-CM: 'N18', 'N19', 'I120', 'I131', 'N250', 'Z490', 'Z491', 'Z492', 'Z940', 'Z992' |
| Liver disease | ICD-10-CM: 'B18', 'I85', 'K70', 'K72', 'K73', 'K74', 'I864', 'I982', 'K711', 'K713', 'K714', 'K715', 'K717', 'K760', 'K762', 'K763', 'K764', 'K765', 'K766', 'K767', 'K768', 'K769', 'Z944' |
| Peptic ulcer disease | ICD-10-CM: 'K257', 'K259', 'K267', 'K269', 'K277', 'K279', 'K287', 'K289' |
| HIV/AIDS | ICD-10-CM: 'B20', 'B21', 'B22', 'B24' |
| Lymphoma | ICD-10-CM: 'C81', 'C82', 'C83', 'C84', 'C85', 'C88', 'C96', 'C900', 'C902' |
| Solid tumor without metastasis | ICD-10-CM: 'C00', 'C01', 'C02', 'C03', 'C04', 'C05', 'C06', 'C07', 'C08', 'C09', 'C10', 'C11', 'C12', 'C13', 'C14', 'C15', 'C16', 'C17', 'C18', 'C19', 'C20', 'C21', 'C22', 'C23', 'C24', 'C25', 'C26', 'C30', 'C31', 'C32', 'C33', 'C34', 'C37', 'C38', 'C39', 'C40', 'C41', 'C43', 'C45', 'C46', 'C47', 'C48', 'C49', 'C50', 'C51', 'C52', 'C53', 'C54', 'C55', 'C56', 'C57', 'C58', 'C60', 'C61', 'C62', 'C63', 'C64', 'C65', 'C66', 'C67', 'C68', 'C69', 'C70', 'C71', 'C72', 'C73', 'C74', 'C75', 'C76', 'C97' |
| Metastatic cancer | ICD-10-CM: 'C77', 'C78', 'C79', 'C80' |
| Rheumatoid arthritis | ICD-10-CM: 'M05', 'M06', 'M08', 'M30', 'M32', 'M33', 'M34', 'M35', 'M45', 'L940', 'L941', 'L943', 'M120', 'M123', 'M310', 'M311', 'M312', 'M313', 'M461', 'M468', 'M469' |
| Coagulopathy | ICD-10-CM: 'D65', 'D66', 'D67', 'D68', 'D691', 'D693', 'D694', 'D695', 'D696' |
| Obesity | ICD-10-CM: 'E66' |
| Weight loss | ICD-10-CM: 'E40', 'E41', 'E42', 'E43', 'E44', 'E45', 'E46', 'R64', 'R634' |
| Fluid and electrolyte disorders | ICD-10-CM: 'E86', 'E87', 'E222' |
| Blood loss anemia | ICD-10-CM: 'D500' |
| Iron deficiency anemia | ICD-10-CM: 'D51', 'D52', 'D53', 'D508', 'D509' |
| Alcohol abuse | ICD-10-CM: 'F10', 'E52', 'T51', 'G621', 'I426', 'K292', 'K700', 'K703', 'K709', 'Z502', 'Z714', 'Z721' |
| Drug abuse | ICD-10-CM: 'F11', 'F12', 'F13', 'F14', 'F15', 'F16', 'F18', 'F19', 'Z715', 'Z722' |
| Psychoses | ICD-10-CM: 'F20', 'F22', 'F23', 'F24', 'F25', 'F28', 'F29', 'F302', 'F312', 'F315' |
| Depression | ICD-10-CM: 'F32', 'F33', 'F204', 'F313', 'F314', 'F315', 'F341', 'F412', 'F432' |
| Sleep disorders | ICD-10-CM: 'G47' |
| Multiple sclerosis | ICD-10-CM: 'G45' |
| Pneumonia | ICD-10-CM: 'J120', 'J121', 'J122', 'J1281', 'J123', 'J1289', 'J129', 'J13', 'J181', 'J150', 'J151', 'J14', 'J154', 'J153', 'J1520', 'J15211', 'J15212', 'J1529', 'J158', 'J155', 'J156', 'A481', 'J159', 'J157', 'J160', 'J168', 'B250', 'A3701', 'A3711', 'A3781', 'A3791', 'A221', 'B440', 'J17', 'B7781', 'J180', 'J188', 'J189', 'J1000', 'J1001', 'J1008', 'J1100', 'J1108', 'J129' |
| Septicemia | ICD-10-CM: 'A021', 'A207', 'A227', 'A312', 'A392', 'A393', 'A394', 'A400', 'A401', 'A403', 'A408', 'A409', 'A412', 'A4101', 'A4102', 'A411', 'A403', 'A414', 'A4150', 'A413', 'A4151', 'A4152', 'A4153', 'A4159', 'A4181', 'A4189', 'A427', 'A267', 'A327', 'A5486', 'B377', 'A419', 'B007', 'R7881', 'A021', 'A227', 'A267', 'A327', 'A400', 'A401', 'A403', 'A408', 'A409', 'A4101', 'A4102', 'A411', 'A412', 'A413', 'A414', 'A4150', |
|  | 'A4151', 'A4152', 'A4153', 'A4159', 'A4181', 'A4189', 'A419', 'A427', 'A5486', 'B377', 'R6520', 'R6521' |
| Stroke | ICD-10-CM: 'G45', 'H341', 'I60', 'I61', 'I63', 'I64' |
| Annual wellness visits | HCPCS: 'G0438', 'G0439' |
| Flu vaccination | HCPCS: '90630', '90653', '90654', '90655', '90656', '90657', '90661', '90662', '90672', '90673', '90674', '90682', '90685', '90686', '90687', '90688', '90756', 'Q2035', 'Q2037', 'Q2038' |
| Chronic care management | HCPCS: '99487', '99489', '99490', '99491', 'G0506' |

